# Artificial intelligence-assisted meta-analysis of the frequency of *ACE* I/D polymorphisms in centenarians and other long-lived individuals

**DOI:** 10.1101/2022.12.10.22283306

**Authors:** Lingxuan Li, Shin Murakami

**Author notes:** Corresponding author, Shin Murakami, PHD, FGSA.

## Abstract

Current research on the angiotensin-converting-enzyme (ACE) gene has yielded controversial results on whether different ACE polymorphisms are linked with human longevity. ACE polymorphisms are a risk factor for Alzheimer’s disease and age-onset diseases that may contribute to the mortality of older people. Our goal is to consolidate existing studies with assistance from artificial intelligence and machine-learning-assisted software to come to a more precise understanding of the role of the ACE gene in human longevity. The I (insertion) and D (deletion) polymorphisms in the intron are correlated with the levels of circulating ACE; homozygous D (DD) is high and homozygous I (II) is low. Here, we performed a detailed meta-analysis of the I and D polymorphisms using centenarians (100+ years old), long-lived subjects (85+ years old), and control groups. ACE genotype distribution was analyzed across a total of 2,054 centenarians and 12,074 controls, as well as 1,367 long-lived subjects between the ages of 85-99, using the inverse variance and random effects methods. The ACE DD genotype was found to be favored in centenarians (OR: 1.41 [95% CI: 1.19-1.67], P < 0.0001) with a heterogeneity of 32%, and the II genotype slightly favored the control groups (OR: 0.81 [95% CI: 0.66-0.98], P = 0.03) with a heterogeneity of 28%, corroborating results from previous meta-analyses. Novel to our meta-analysis, the ID genotype was found to be favored in control groups (OR: 0.86 [95% CI: 0.76-0.97], P = 0.01) with a heterogeneity of 0%. The long-lived group showed a similar positive association between the DD genotype and longevity (OR: 1.34 [95% CI: 1.21-1.48, P < 0.0001) and a negative association between the II genotype and longevity (OR: 0.79 [95% CI: 0.70-0.88], P < 0.0001). The long-lived ID genotype did not show significant findings (OR: 0.93 [95% CI: 0.84-1.02], P = 0.79). In conclusion, the results suggest a significant positive association of the DD genotype with human longevity. However, despite the previous study, the results do not confirm a positive association of the ID genotype with human longevity. We suggest a few paradoxical observations: (1) inhibition of ACE can increase longevity in model systems from nematodes to mammals, seemingly opposite to the finding in humans; (2) exceptional longevity associated with homozygous DD is also associated age-related diseases with higher mortality risks in homozygous DD. We discuss ACE, longevity, and age-related diseases.

## Introduction

Human life expectancy has increased in the past millennia since the studies of longevity in ancient Greek and Roman populations (Montagu, 1994; Batrinos, 2008; Roser et al., 2013). The study of aging, including the biology of aging and populations of older people, has suggested that longevity is influenced by a combination of genetic, lifestyle, and environmental factors (reviewed in Weinert et al., 2003). The search for genetic causes of longevity has implicated several possible candidate genes, including SIRT1, APOE, FOXO3A, ACE, ATM, NOS1, NOS2, KLOTHO and IL6 (reviewed in Revelas et al., 2018; Costa et al., 2019). Of them, ACE encodes an angiotensin-converting enzyme, which has two functions with opposing effects on health (Duc et al., 2020; Duc et al., 2021). Firstly, ACE is an essential enzyme for hemodynamic control via the renin-angiotensin-aldosterone system and the kinin-kallikrein system. ACE converts the inactive angiotensin I to the active angiotensin II, a potent vasoconstrictor, and inactivates bradykinin, a potent vasodilator. ACE inhibitors are one of the first-line treatments for hypertension and congestive heart failure. In this function, ACE inhibition is beneficial to health through controlling hypertension. Secondly, ACE is an amyloid-degrading enzyme that can reduce the proteo-toxicity of amyloid. Importantly, ACE, when altered, is a risk factor gene in Alzheimer’s disease (Vahdati Nia et al., 2017; Murakami and Lacayo, 2022). ACE alterations are associated with Alzheimer’s disease (discussed in Duc et al., 2021). In this function, ACE inhibition may be deleterious to the health through amyloid toxicity. Thus, the effect of ACE on longevity may differ depending on the health conditions of advanced aging (Duc et al., 2021). Thus, we reason that long-lived groups should be included in this study.

The ACE gene is located on chromosome 17q23. The ACE I/D polymorphisms were first identified in 1990, which are characterized by the presence (insertion, I) or absence (deletion, D) of a 287-bp (base pair) Alu repeat sequence in intron 16 (Rigat et al., 1990). The I/D polymorphisms correlated with the serum levels of circulating ACE; the D-allele is higher and the I-allele is lower (Tiret et al., 1992). The DD individuals have approximately twice the plasma ACE levels as the II individuals do (Rigat et al., 1990; Tomita et al., 1996). Other meta-analyses have also shown a positive association between the D-allele and increased risk of essential hypertension (Li et al., 2011), ischemic stroke (Zhang et al., 2012), coronary artery disease (Zintzaras et al., 2008), left ventricular hypertrophy (Li et al., 2012) and pneumonia (Nie et al., 2014). In addition, the D-allele has been linked with higher mortality rates from COVID-19 (Yamamoto et al., 2020).

Paradoxically, the ACE D allele is associated with the risk of cardiovascular disease, hypertension, Alzheimer’s disease, and other common causes of death in older people, while a previous study noted an increased frequency of the DD genotype among a group of centenarians from France (Schachter et al., 1994). Since then, centenarian studies have been performed across different cohorts, resulting in conflicting results. A more recent study suggests that the DD genotype and the D allele are enriched in centenarians (Garatachea et al., 2013), while another study suggests that no significant difference among the I and D alleles (Choi et a., 2003; Oscanoa, et al., 2020). In this meta-analysis, we further consolidated these findings to more precisely determine whether or not the D polymorphism is enriched in centenarians and long-lived individuals.

## Methods

### Nomenclature, PRISMA and Database search

Due to nomenclature inconsistency among studies, we used I (Insertion) and D (deletion) alleles as follows: The genotypes were referred to as homozygous insertion (II), heterozygous insertion/deletion (ID), and homozygous deletion (DD). The I and D polymorphisms were referred to as I/D. We followed the Preferred Reporting Items for Systematic Reviews and Meta-Analyses (PRISMA) (Moher et al., 2009) and performed a meta-analysis as described (Antos et al., 2021) with updates described in PRISMA 2020 (Page et al., 2021). We performed a literature search to collect all publications on ACE I/D polymorphisms in centenarians published before May 25, 2022. A total of 80 studies were obtained from the Human Genomic Aging Resource’s Longevity Map database, Scopus, PubMed, and Google Scholar using the keywords “ACE” OR “angiotensin-converting enzyme” AND “longevity” OR “centenarian” OR “nonagenarian” (subjects 90+ years). Studies of all languages were included. The titles and abstracts of each article were imported to Colandr (Cheng et al., 2018) which uses AI and machine-learning algorithms to determine an article’s relevance based on text-based evidence. Of the 80 studies initially identified, 37 were excluded as duplicates. The remaining 43 articles were then screened in Colandr according to specified inclusion criteria.

### Inclusion criteria

Studies were included if they met all of the following conditions: (1) evaluation of the ACE I/D polymorphisms and longevity; (2) case-control study design; (3) study provides sample size, genotype data, odds ratio (OR) and 95% confidence interval (CI), or the data necessary to calculate the results. The exclusion criteria of the studies were as follows: (1) not relevant to ACE I/D polymorphisms or longevity; (2) review or meta-analysis; (3) minimum long-lived population age < 85 years or unspecified; (4) no control group or maximum control group age > 85 years. Data was then separated into centenarian-only and long-lived (85+ years old) sections. Studies were included in the centenarian-only section if the mean population age was > 100. In total, we obtained 13 papers that met all our inclusion criteria for centenarians and 19 papers for the long-lived group.

### Data extraction

Data were extracted independently by the author and an assistant, and any discrepancies were settled by consensus. The data extracted included the author’s name, year of publication, ethnicity of the study population, sample size, and genotype numbers in long-lived and control groups. Allele frequencies were calculated if not directly reported. If the study had multiple control groups of varying age ranges, the control group was defined as all individuals ≤ 85 years of age, and data was extracted accordingly.

### Data synthesis and assessment of the risk of bias

To determine the association between ACE genotype and longevity, the odds ratio (OR) with the corresponding 95% confidence interval (CI) was calculated for each genotype (DD, ID, and II). All statistical analyses were performed using The Cochrane Collaboration’s Review Manager (RevMan) Version 5.4 using the random effects statistical model and inverse variance methods. The risk of bias in the studies was performed as described by Antos et al., 2021. The result was visualized with Risk-of-bias VISualization (robvis) ROBINS-1 (Sterne et al., 2016; McGuinness et al., 2021). Bias due to deviations from intended interventions did not apply to this meta-analysis, as participants were categorized by age and could not be blinded in which group they were assigned.

## Results

### I. Centenarian results

Our literature search is summarized in Figure 1. The extracted data of centenarians from the identified studies are summarized in Table 1. The meta-analysis included 13 studies (Bladbjerg et al., 1999; Blanche et al., 2001; Choi et al., 2003; Pereira da Silva et al., 2018; Faure-Delanef et al., 1998; Fiuza-Luces et al., 2011; Nacmias et al., 2007; Panza et al., 2002, 2003; Schachter et al., 1994; Seripa et al., 2006; Wufuer et al., 2004; Zhang et al., 2010), involving a total of 2,054 centenarians and 10,986 controls. Ethnicities of the studied populations were Caucasian (n = 10), Korean (n = 1), Uyghur (n = 1) and Han Chinese (n = 1). This meta-analysis investigated what age groups could be analyzed. We found three age groups that consistently showed up among the previous studies, resulting in centenarians (100+ years old), long-lived groups (85+ years old), and control groups. The range of the control groups was diverse among previous studies. We set the ages of control groups as 18-85 year old which was judged to be inclusive to most of the studies.

**Table 1.**
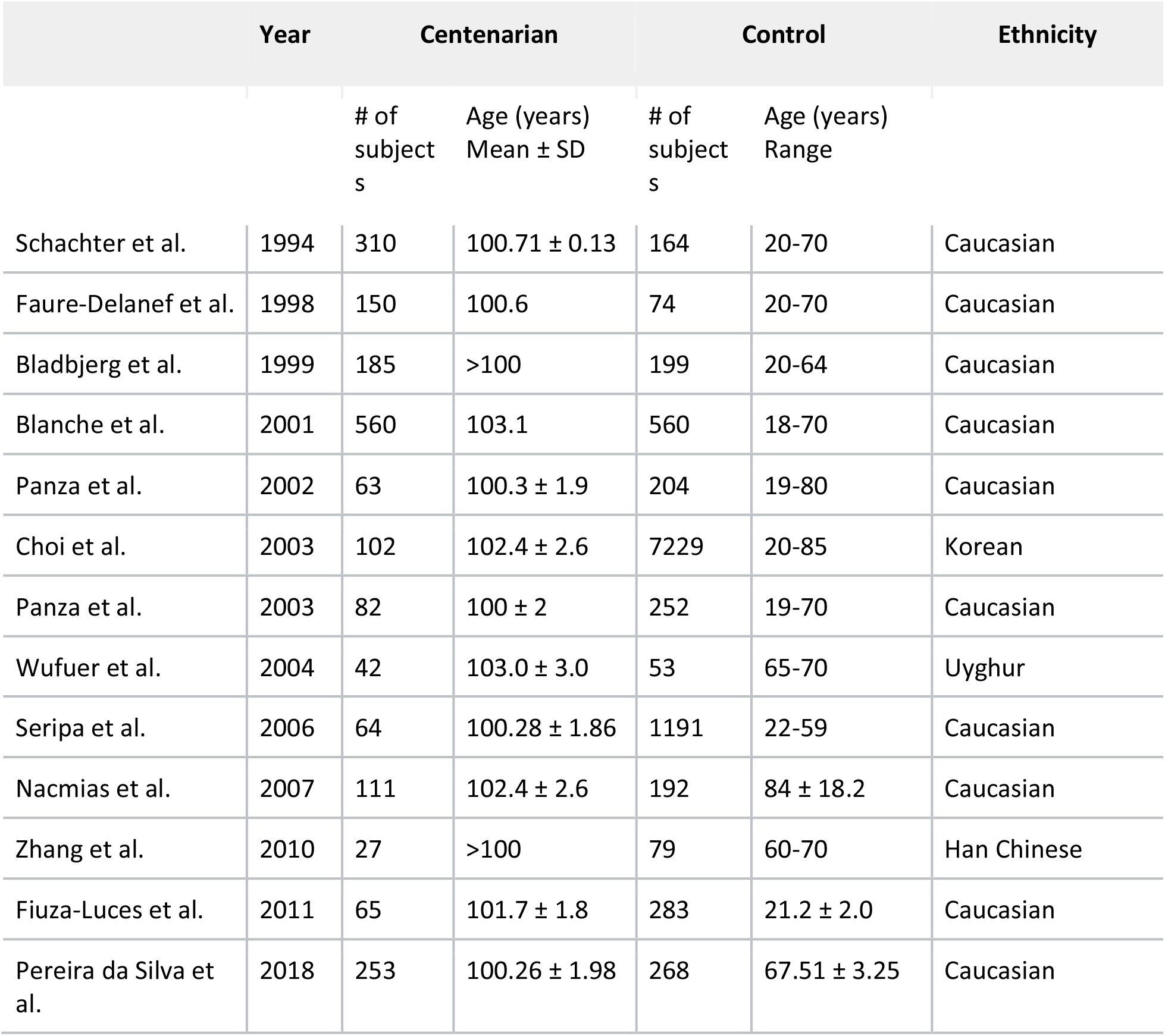
Characteristics of studies on *ACE* I/D polymorphisms in centenarians. A total of 13 studies were included that looked at the frequency of different *ACE* I/D polymorphisms in centenarians.

|  | Year | Centenarian |  | Control |  | Ethnicity |
| --- | --- | --- | --- | --- | --- | --- |
| | | # of subjects | Age (years)<br>Mean ± SD | # of subjects | Age (years)<br>Range | |
| Schachter et al. | 1994 | 310 | 100.71 ± 0.13 | 164 | 20-70 | Caucasian |
| Faure-Delanef et al. | 1998 | 150 | 100.6 | 74 | 20-70 | Caucasian |
| Bladbjerg et al. | 1999 | 185 | >100 | 199 | 20-64 | Caucasian |
| Blanche et al. | 2001 | 560 | 103.1 | 560 | 18-70 | Caucasian |
| Panza et al. | 2002 | 63 | 100.3 ± 1.9 | 204 | 19-80 | Caucasian |
| Choi et al. | 2003 | 102 | 102.4 ± 2.6 | 7229 | 20-85 | Korean |
| Panza et al. | 2003 | 82 | 100 ± 2 | 252 | 19-70 | Caucasian |
| Wufuer et al. | 2004 | 42 | 103.0 ± 3.0 | 53 | 65-70 | Uyghur |
| Seripa et al. | 2006 | 64 | 100.28 ± 1.86 | 1191 | 22-59 | Caucasian |
| Nacmias et al. | 2007 | 111 | 102.4 ± 2.6 | 192 | 84 ± 18.2 | Caucasian |
| Zhang et al. | 2010 | 27 | >100 | 79 | 60-70 | Han Chinese |
| Fiuza-Luces et al. | 2011 | 65 | 101.7 ± 1.8 | 283 | 21.2 ± 2.0 | Caucasian |
| Pereira da Silva et al. | 2018 | 253 | 100.26 ± 1.98 | 268 | 67.51 ± 3.25 | Caucasian |

**Figure 1.**
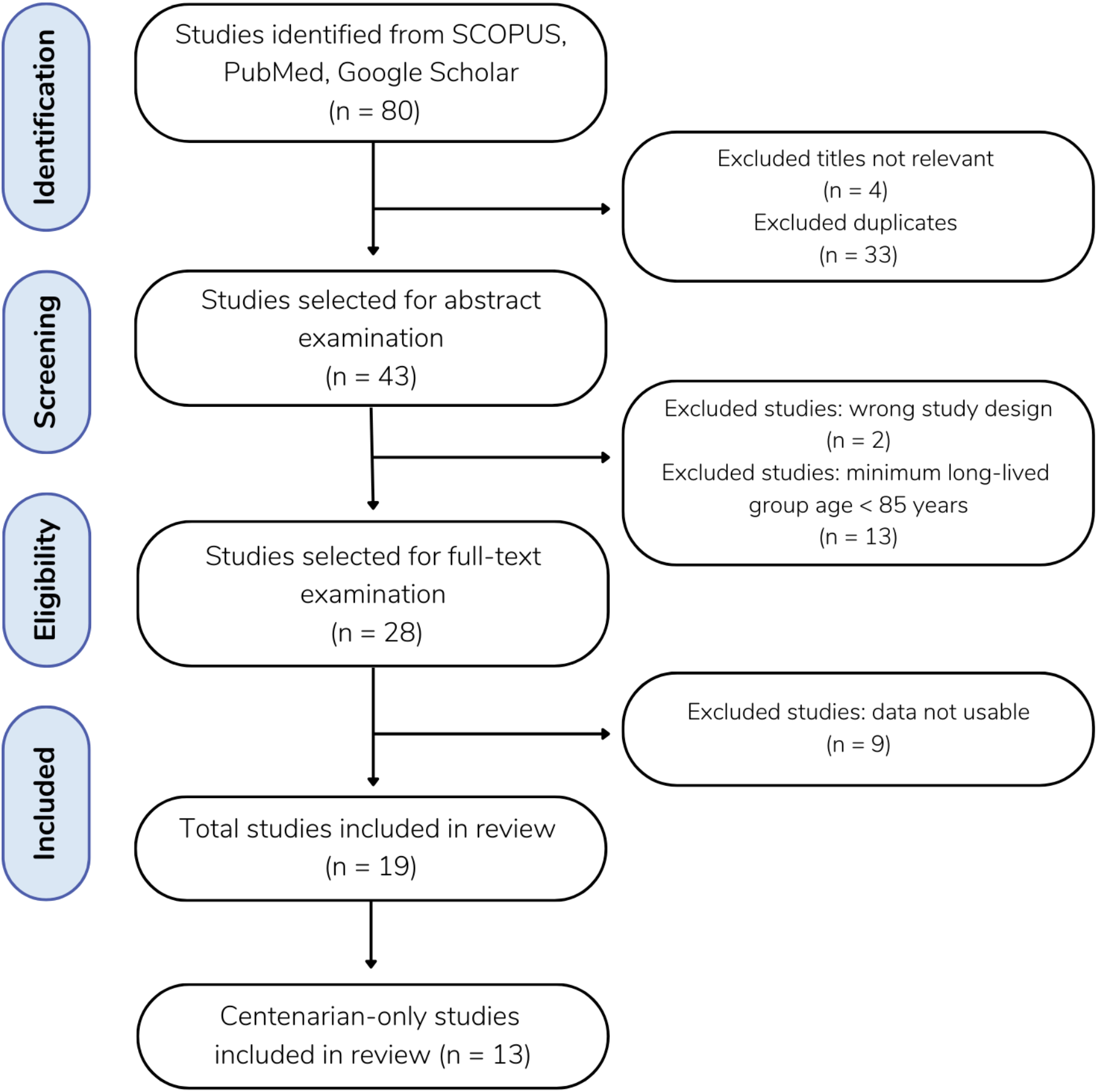
Flow diagram of the literature search. A total of 80 studies were gathered from SCOPUS (n = 39), PubMed (n = 26) and Google Scholar (n = 15). Excluding 33 duplicates and 4 studies with irrelevant titles, 43 remaining studies were selected for abstract examination. Following the abstract screening, 2 studies were excluded for having the wrong study design and 13 studies were excluded for defining their minimum long-lived population age to be < 85 years of age. The full-text review was performed on the remaining 28 studies, and 9 of those were excluded as the data was not usable, meaning that the study collected and presented data in a form other than the frequency of individuals genotypes and that the frequency of genotypes was not able to be calculated from the data available.

The major differences from the previous meta-analysis were as follows. Firstly, we used different age groups for a more precise understanding of the effect of ACE on longevity. The previous meta-analysis compared the frequencies of various combinations of I and D (i.e., DD vs. II; DD vs. ID; ID vs. II; DD + ID vs. II; and DD vs. ID + II) (Garatachea et al., 2013), which lacked comparisons among different age groups. Secondly, we excluded a study (Paolisso et al., 2001), which was used in the previous meta-analysis (Garatachea et al., 2013) for the following reason: The excluded study used 62-88 years old, which overlapped with the age range for the long-lived group (85+ years old) commonly used in previous studies (see II. Long-Lived Results). Finally, we incorporated AI-assisted software to reduce the burden of the meta-analysis studies.

As shown in Figure 2, the association between the DD genotype and exceptional longevity was analyzed in 13 studies (Bladbjerg et al., 1999; Blanche et al., 2001; Choi et al., 2003; Pereira da Silva et al., 2018; Faure-Delanef et al., 1998; Fiuza-Luces et al., 2011; Nacmias et al., 2007; Panza et al., 2002, 2003; Schachter et al., 1994; Seripa et al., 2006; Wufuer et al., 2004; Zhang et al., 2010). The DD genotype was more frequent among centenarians than in controls, with an OR of 1.41 (95% CI: 1.19-1.67, P < 0.0001) with mild heterogeneity (32%), indicating a significant positive association between the DD genotype and longevity.

**Figure 2:**
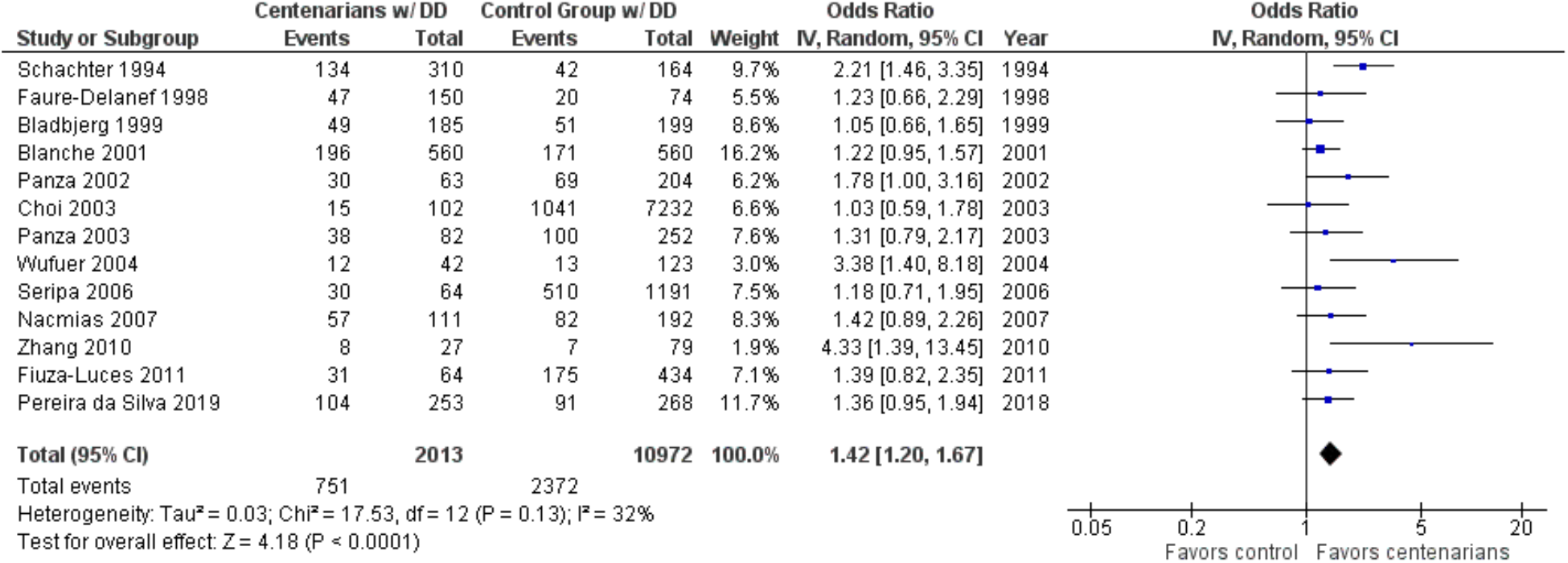
Comparison of DD genotype frequency in centenarians and control groups. The ages of centenarians and control groups are as described in Table 1.

As shown in Figure 3, the association between the ID genotype and exceptional longevity was analyzed in 13 studies (Bladbjerg et al., 1999; Blanche et al., 2001; Choi et al., 2003; Pereira da Silva et al., 2018; Faure-Delanef et al., 1998; Fiuza-Luces et al., 2011; Nacmias et al., 2007; Panza et al., 2002, 2003; Schachter et al., 1994; Seripa et al., 2006; Wufuer et al., 2004; Zhang et al., 2010). The ID genotype was found to have an OR of 0.86 (95% CI: 0.76-0.97, P = 0.01) with no evidence of heterogeneity (0%), indicating a significant negative association between the ID genotype and longevity.

**Figure 3:**
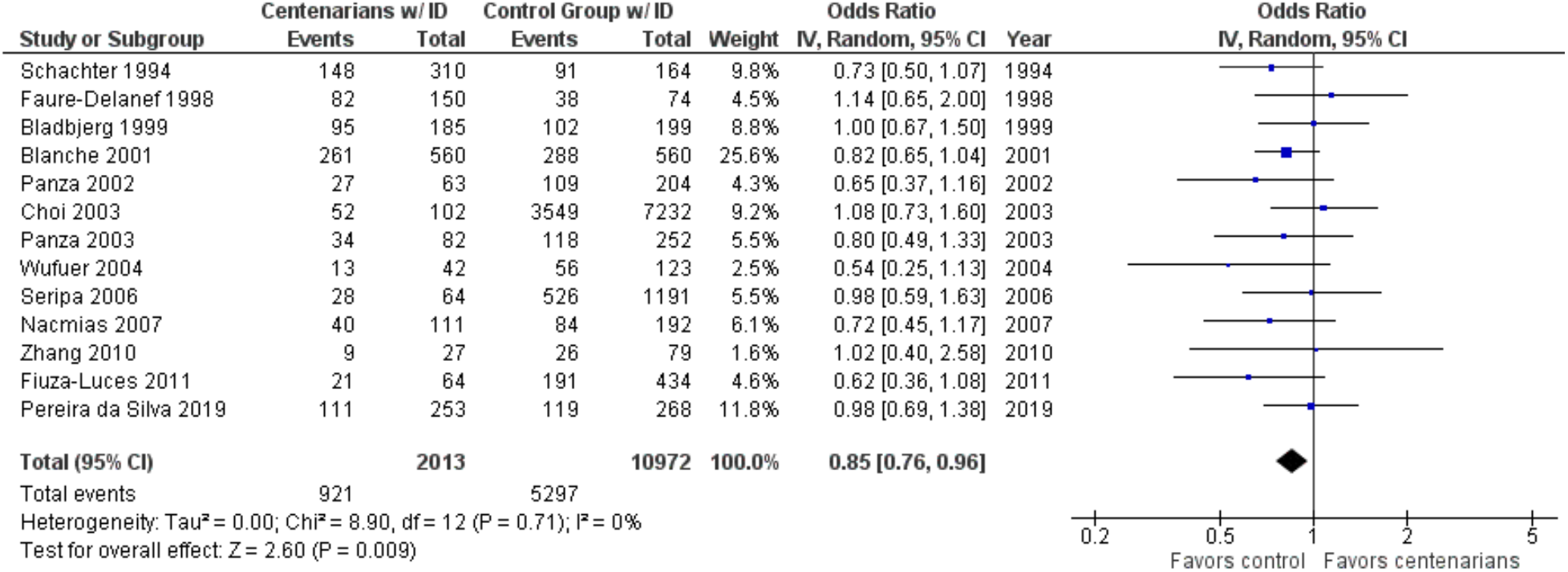
Comparison of ID genotype frequency in centenarians and control groups. The ages of centenarians and control groups are described in Table 1.

As shown in Figure 4, the association between the II genotype and exceptional longevity was analyzed in 13 studies (Bladbjerg et al., 1999; Blanche et al., 2001; Choi et al., 2003; Pereira da Silva et al., 2018; Faure-Delanef et al., 1998; Fiuza-Luces et al., 2011; Nacmias et al., 2007; Panza et al., 2002, 2003; Schachter et al., 1994; Seripa et al., 2006; Wufuer et al., 2004; Zhang et al., 2010). The II genotype was found to have an OR of 0.81 (95% CI: 0.66-0.98, P = 0.03) with mild heterogeneity (28%), indicating a significant negative association between the II genotype and longevity.

**Figure 4:**
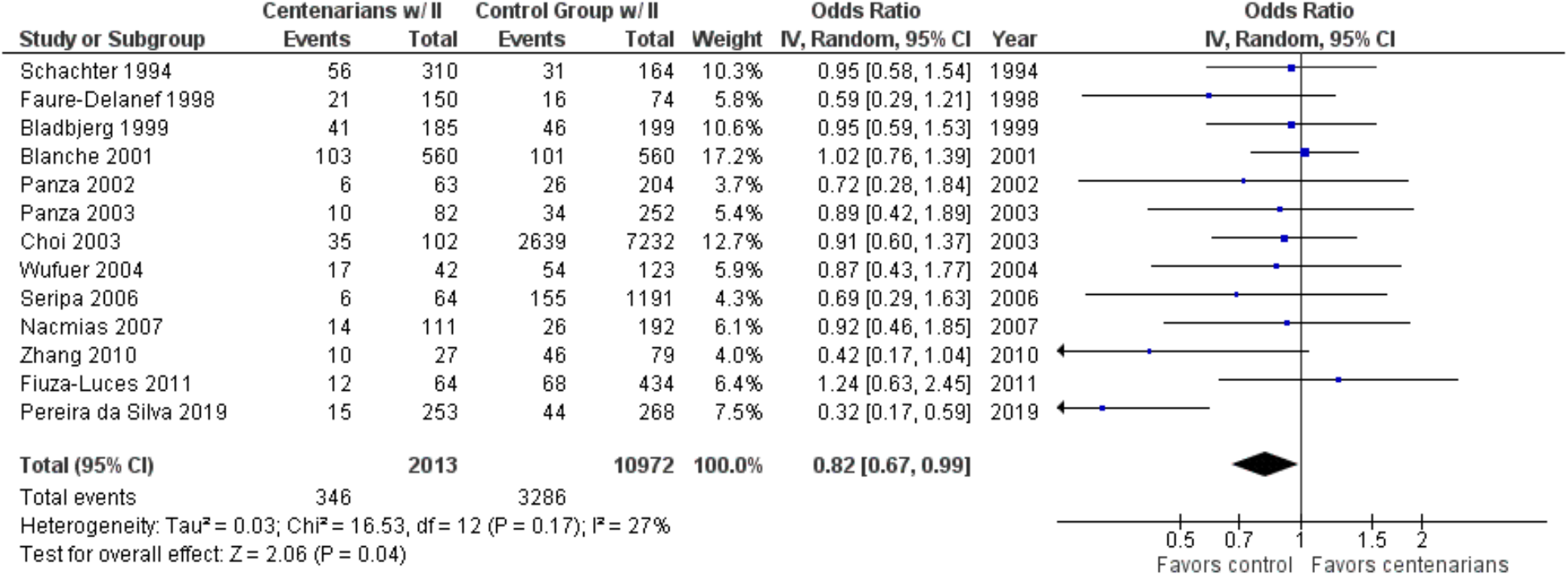
Comparison of II genotype frequency in centenarians and control groups. The ages of centenarians and control groups are as described in Table 1.

### II. Long-Lived Results

Table 2 summarizes the data of long-lived groups. The meta-analysis included 19 studies, with 13 centenarian-only studies, and 5 studies that included both nonagenarians and centenarians (Arkhipova et al., 2014; Kolovou et al., 2014; Lo Sasso et al., 2012; Oscanoa et al., 2020; Yang et al., 2009), and 1 study with individuals 85 years and older (Heijmans et al., 1999), involving a total of 3,421 long-lived individuals (85+ years old) and 11,959 controls. Ethnicities of the studied populations were Caucasian (n = 13), Korean (n = 1), Uyghur (n = 1), Han Chinese (n = 2), Russian/Yakut (n = 1), and Peruvian (n = 1).

**Table 2.**
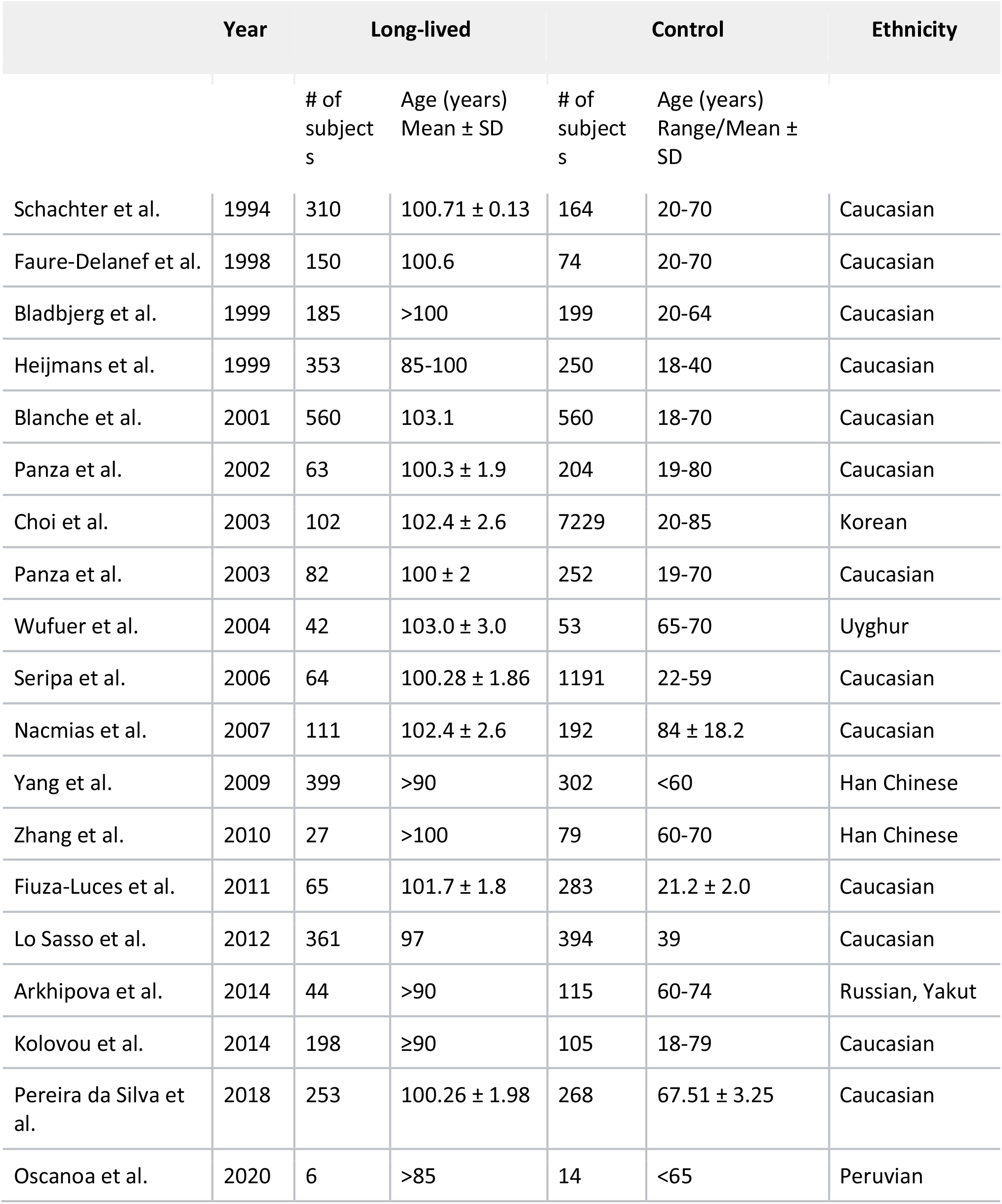
Characteristics of studies on *ACE* I/D polymorphisms in long-lived individuals (85+ years old).

The association between the DD genotype and exceptional longevity was analyzed in 18 studies (Bladbjerg et al., 1999; Blanche et al., 2001; Choi et al., 2003; Pereira da Silva et al., 2018; Faure-Delanef et al., 1998; Fiuza-Luces et al., 2011; Kolovou et al., 2014; Lo Sasso et al., 2012; Nacmias et al., 2007; Oscanoa et al., 2020; Panza et al., 2002, 2003; Schachter et al., 1994; Seripa et al., 2006; Wufuer et al., 2004; Yang et al., 2009; Zhang et al., 2010). The DD genotype was found to have an OR of 1.32 (95% CI: 1.19-1.47, P < 0.00001) with mild heterogeneity (30%), indicating a significant positive association between the DD genotype and longevity.

As shown in Figure 6, the association between the ID genotype and exceptional longevity was analyzed in 18 studies (Bladbjerg et al., 1999; Blanche et al., 2001; Choi et al., 2003; Pereira da Silva et al., 2018; Faure-Delanef et al., 1998; Fiuza-Luces et al., 2011; Kolovou et al., 2014; Lo Sasso et al., 2012; Nacmias et al., 2007; Oscanoa et al., 2020; Panza et al., 2002, 2003; Schachter et al., 1994; Seripa et al., 2006; Wufuer et al., 2004; Yang et al., 2009; Zhang et al., 2010).

**Figure 5.**
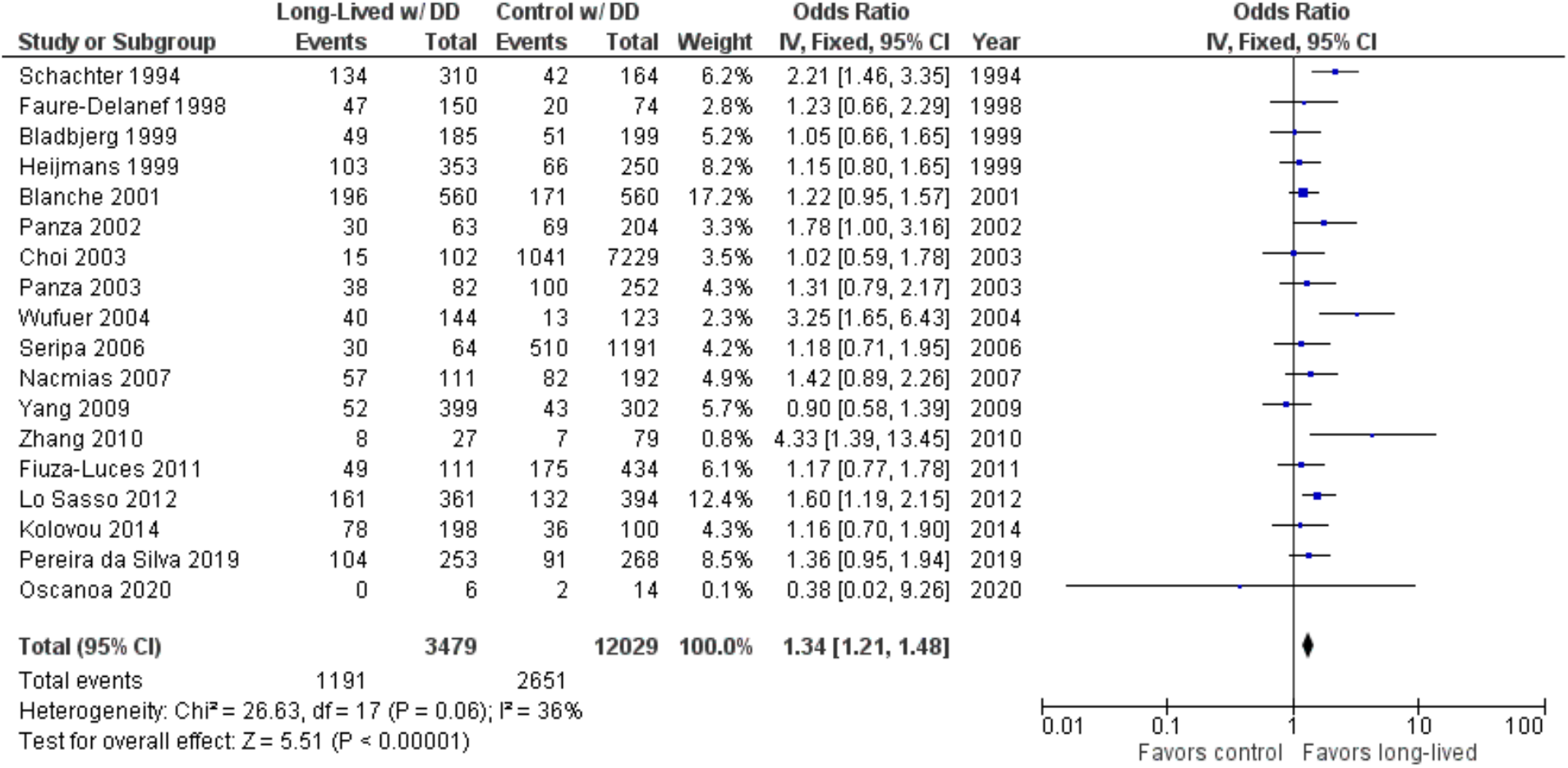
Comparison of DD genotype frequency in long-lived and control groups. The ages of long-lived and control groups are described in Table 1.

**Figure 6.**
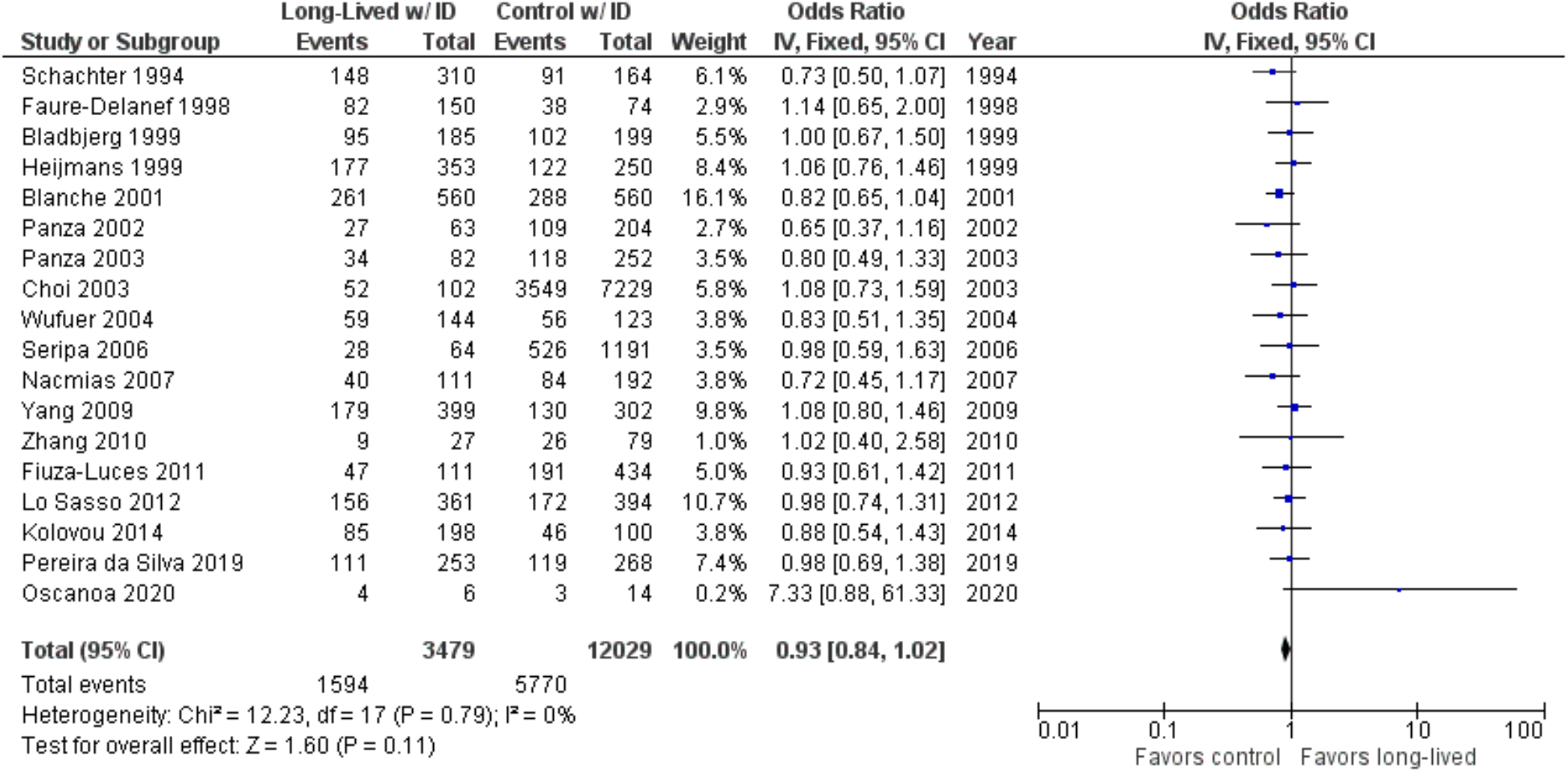
Comparison of ID genotype frequency in long-lived and control groups. The ages of long-lived and control groups are described in Table 1.

The ID genotype was found to have an OR of 0.92 (95% CI: 0.84-1.02, P = 0.11) with no evidence of heterogeneity (0%). There was no significant difference in the frequency of the ID genotype between the long-lived (85+ years old) and control groups.

As shown in Figure 7, the association between the II genotype and exceptional longevity was analyzed in 19 studies (Arkhipova et al., 2014; Bladbjerg et al., 1999; Blanche et al., 2001; Choi et al., 2003; Pereira da Silva et al., 2018; Faure-Delanef et al., 1998; Fiuza-Luces et al., 2011; Kolovou et al., 2014; Lo Sasso et al., 2012; Nacmias et al., 2007; Oscanoa et al., 2020; Panza et al., 2002, 2003; Schachter et al., 1994; Seripa et al., 2006; Wufuer et al., 2004; Yang et al., 2009; Zhang et al., 2010). Arkhipova et al., 2014 only provided raw frequency data for the II genotype and thus was not included in the DD or ID analyses. The II genotype was found to have an OR of 0.80 (95% CI: 0.71-0.90, P = 0.002) with mild heterogeneity (30%), indicating a significant negative association between the II genotype and longevity. The risk of bias of the studies was assessed using ROBINS-1 (Method). As shown in Figure 8, the result suggests an overall low risk of bias.

**Figure 7.**
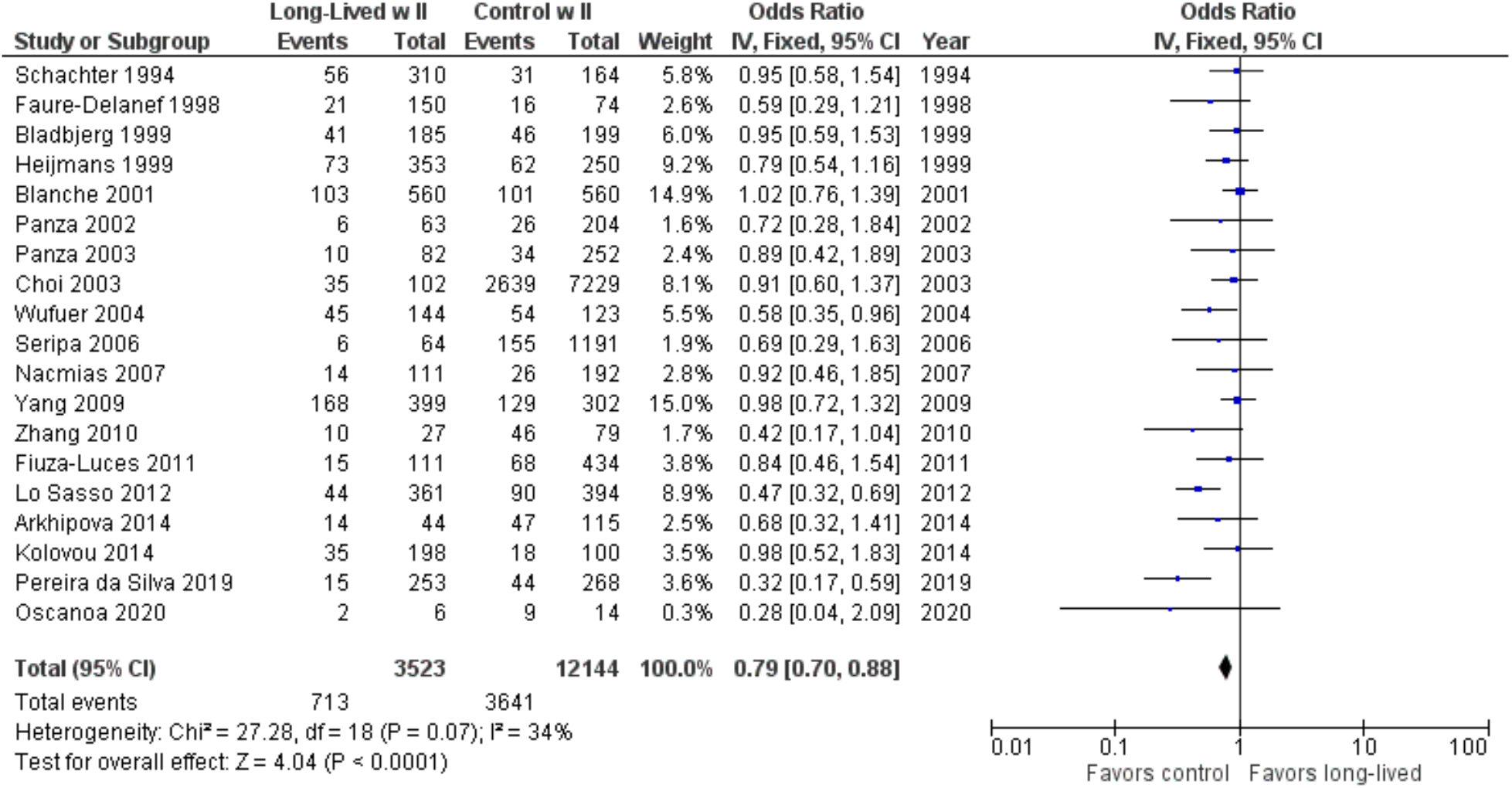
Comparison of II genotype frequency in long-lived and control groups. The ages of long-lived and control groups are described in Table 1.

**Figure 8A.**
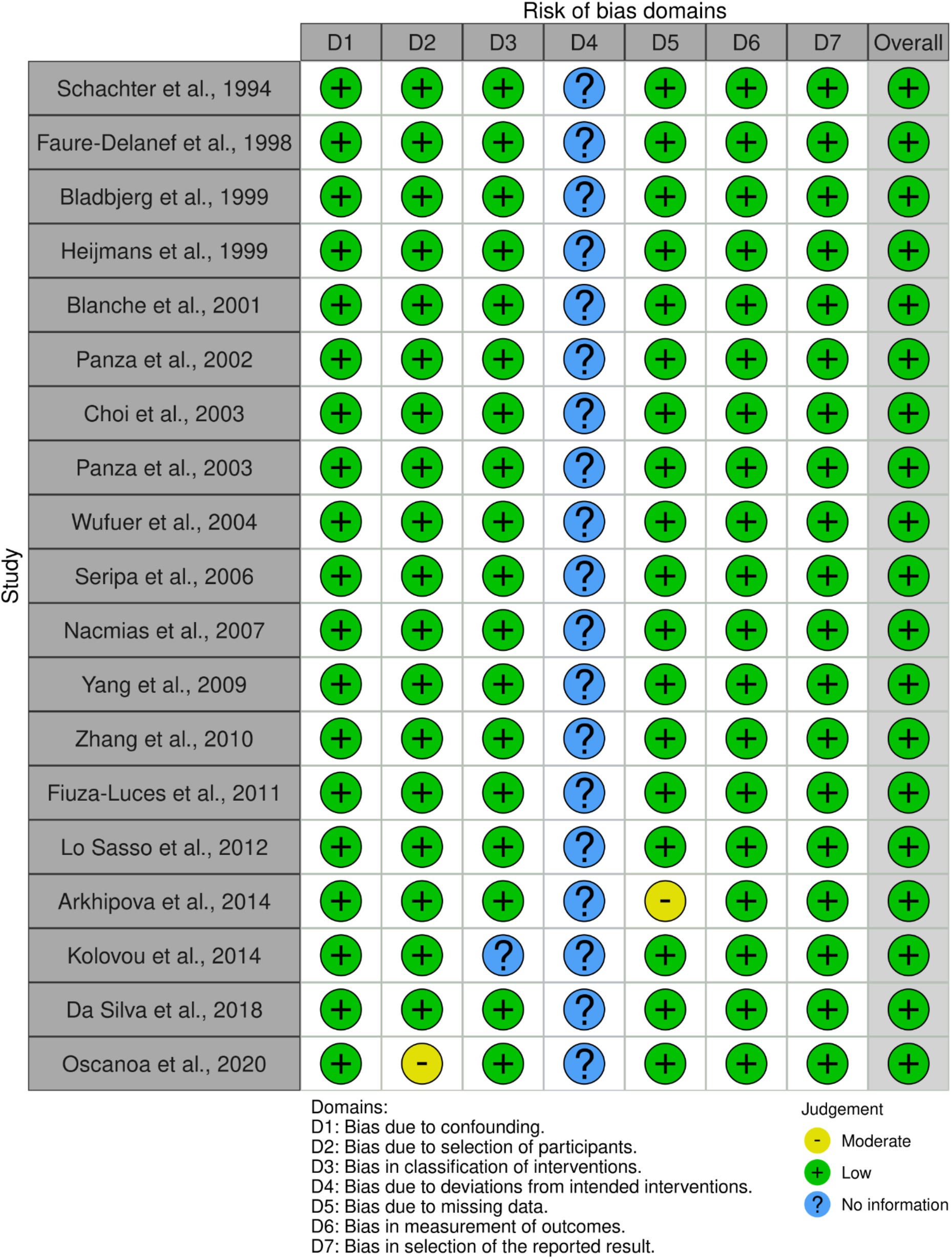
Traffic light plot showing the assessment of the risk of bias using ROBINS-1.

**Figure 8B.**
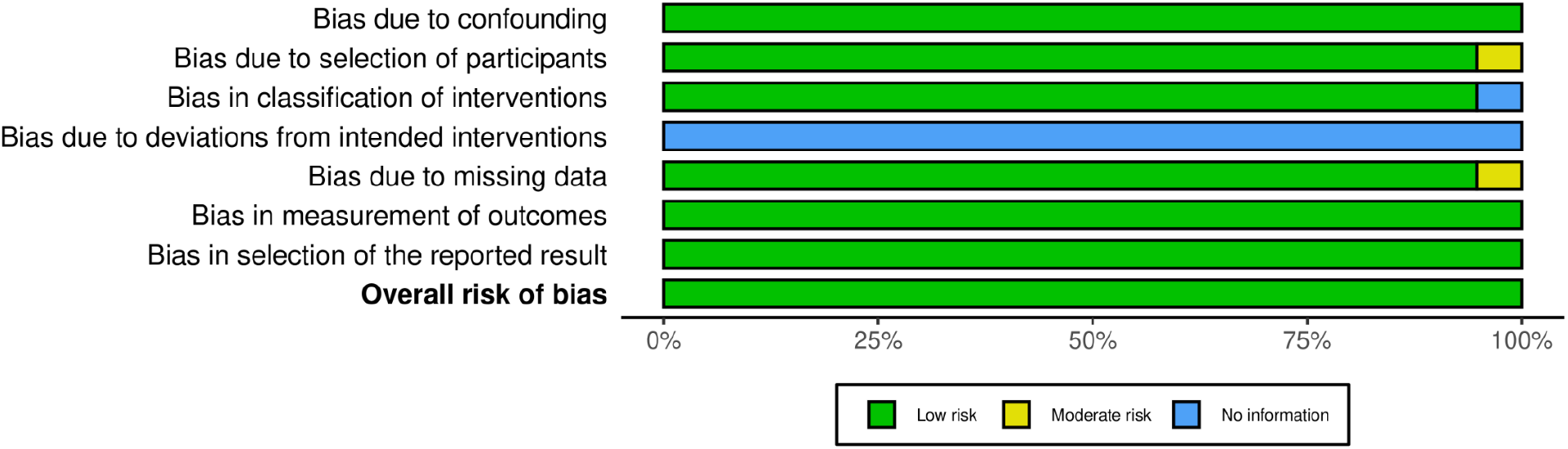
Summary plot showing the assessment of the risk of bias using ROBINS-1.

## Discussion

Centenarians are the fastest-growing demographic group of the world’s population, having roughly doubled every decade since 1950 (Wilcox et al., 2011). It has been nearly a decade since the most recent meta-analysis has been published in 2013 (Garatachea et al., 2013) and additional studies on centenarians and nonagenarians have been conducted that further elucidate the association between *ACE* genotype and longevity. In addition, the use of AI and machine-learning software has aided in the search and screening process to compile even more studies and process the associated data. Our meta-analysis suggests, with increased confidence, that there is a significant positive association of the DD genotype with exceptional longevity, across populations of different ethnicities. It has been reported that serum ACE levels correlate in the order of DD > ID > II in which DD is the highest (Rigat et al., 1990; Tiret et al., 1992; Tomita et al., 1996). The higher levels of ACE were correlated with increased lifespans, especially more in centenarians than in long-lived controls.

The results present a couple of paradoxes in which the D-allele has been reported as a risk factor for hypertension (Li et al., 2011), CVD (Zintzaras et al., 2008), pneumonia (Nie et al., 2014) and COVID-19 (Yamamoto et al., 2020), all of which are major causes of death in older people. However, the DD genotype has been associated with a decreased risk of Alzheimer’s disease, potentially due to ACE’s ability to inhibit amyloid beta aggregation and degrade A beta- (1-40) (Hu et al., 2001; Kölsch et al., 2005; Lehmann et al., 2005). Although the protective effect of the D-allele against Alzheimer’s disease does not fully explain the higher proportion of DD individuals in centenarian cohorts, the D-allele or DD genotype may impart a health risk while also providing a long-term longevity advantage. The middle-life crisis theory of aging (Murakami et al., 2011; Murakami, 2013; Machino et al., 2014; Le et al., 2021), in which midlife events are the catalyst for the transition from normal aging to a more advanced pathological state, can be used to explain ACE’s dual functions as an angiotensin-converting enzyme and an amyloid-degrading enzyme (Le et al., 2021); low levels of ACE plasma activity, associated with the II genotype, may initially confer stress resistance before midlife, but will be insufficient to counter the abnormal accumulation of beta-amyloid when it begins and thus lead to higher mortality rates earlier in the aging process due to AD, while DD individuals who had survived midlife comorbidities would now have lower mortality rates from AD. However, this does not fully explain the ACE paradox, as Alzheimer’s disease is only one of many diseases that affect older people. As the full role of ACE in the human body has not been determined, the D-allele may have some neuroendocrine or immunomodulatory functions that have yet to be discovered (Costerousse et al., 1993) that may further explain ACE’s role in longevity. It raises an unanswered key question how exceptional longevity is associated with positive risks of age-onset diseases that may increase the risk of mortality.

Another paradox is that inhibition of ACE by mutations or drugs can extend lifespans in model systems from nematodes, fruit flies, and mammals (reviewed in Le et al., 2021; Egan et al., 2022). The observation raises awareness that life-extension interventions in model systems may be different from exceptional longevity in humans. Thus, it raises another unanswered key question of whether or not exceptional longevity in humans shares common underlying mechanisms with life extension in model systems.

Our meta-analysis includes a comprehensive review of studies done on centenarians, from Schachter’s initial study in 1994 up to the present, for a total of 2,054 centenarians. We did not limit the literature search to only studies written in English, thus our data represents all of the current *ACE* centenarian studies in the databases searched. The results corroborated the positive association between the DD genotype and exceptional longevity and the negative association between the II genotype and exceptional longevity that had been noted in previous meta-analyses. A novel finding was that the ID genotype, which had previously shown no significant associations with longevity, was found to have a significant negative association with longevity in the centenarian-only analysis.

It is implicated that the effect of ACE on health may differ among different age groups (Duc et al., 2021). Thus, we compared the centenarian-only data with the long-lived (85+ years old) data. We found that the distribution of ACE DD and II genotypes were consistent across both age groups, with strong associations in the DD genotype with exceptional longevity and with negative associations in the II genotype. Interestingly, the ID genotype had a significant negative association among centenarians but did not show a significant association among long-lived groups. The straightforward interpretations are as follows: (1) The DD genotype is associated with centenarians and is long-lived, while the D alleles follow the inheritance pattern of autosomal recessive; (2) The II genotype is negatively associated with longevity in both age groups; (3) The ID genotype shows a weakly negative association with centenarians, while the trend was not significant in long-lived control. Thus, more detailed studies with a series of ages will be needed.

The limitations of this study are as follows. Firstly, it can be summarized as a general limitation of the meta-analysis method. Each search engine list the papers which are indexed by its criteria, which may miss relevant studies. We included three search engines to reduce this risk of missing studies. Secondly, related to the first limitation is that the criteria to analyze the data rely on the criteria used in each study. For example, control groups of the past studies ranged broadly and diverse (18-85 years old). There was a lack of detailed age groups to compare the effect of ACE. Thus, we were not able to have a series of ages to define age-dependent enrichment of the ACE D and I alleles. Thirdly, another limitation is that the vast majority of the centenarian studies on ACE used Caucasian populations. Selectively analyzing the studies of Caucasian populations gives the same results as the pooled data, indicating that there were no major differences in association across ethnicity within this present review. However, there were not enough Asian studies to sufficiently analyze the strength of association within Asian populations only. We also did not find any studies on ACE polymorphisms in Hispanic/Latino or Black centenarian cohorts in our literature search. For future meta-analyses, a greater diversity of such studies should be taken to better illuminate the role of ACE in aging populations across all different environmental or lifestyle risk factors.

## Conclusions

Our meta-analysis suggests a significant positive association between the ACE DD genotype with centenarians and long-lived groups. Although ACE polymorphisms are a risk factor for Alzheimer’s disease and age-onset diseases that may contribute to mortality, the ACE DD genotype but not the ID genotype was in favor of exceptional longevity over the mortality expected in older people. The DD genotype is associated with a reduced risk of Alzheimer’s disease and increased longevity. It is consistent with an ACE function as an amyloid degradation enzyme, which may be important in exceptional longevity in humans. Additionally, the DD genotype causes a high circulating ACE. In contrast, the DD genotype is also associated with an increased risk of various other diseases. They include hypertension (Li et al., 2011), ischemic stroke (Zhang et al., 2012), coronary artery disease (Zintzaras et al., 2008), left ventricular hypertrophy (Li et al., 2012) and pneumonia (Nie et al., 2014). Some of these diseases, including hypertension and cardiovascular conditions (stroke and coronary artery disease), are associated with the risks of COVID-19 mortality (Antos et al., 2021). The DD genotype is associated with higher COVID-19 mortality (Yamamoto et al., 2020). It may imply disease interactions mediated by the D allele. The underlying mechanisms of human longevity remain unclear, especially when considering the D-allele’s role as a risk factor for many age-related comorbidities. Further studies on nonagenarians and centenarians of different backgrounds may provide better insight into the role of ACE as a longevity gene.

## Data Availability

All data produced in the present work are contained in the manuscript

## Author Contributions

L.L. was involved in data collection, organization of AI components, data analysis and writing of the manuscript. S.M. directed and guided all the research process.

## Institutional Review Board Statement

Not applicable.

## Informed Consent Statement

Not applicable.

## Data Availability Statement

The data supporting the reported results have been made available within the manuscript.

